# COVID-19 OUTCOMES IN PREGNANCY: A REVIEW OF 275 SCREENED STUDIES

**DOI:** 10.1101/2021.08.28.21262778

**Authors:** Rupalakshmi Vijayan, Hanna Moon, Jasmine Joseph, Madiha Zaidi, Chhaya Kamwal, Andrelle Senatus, Shavy Nagpal, Miguel Diaz

## Abstract

In December 2019, a novel strain of severe acute respiratory syndrome (SARS-CoV-2), was declared as a cause of respiratory illness, called coronavirus 2019 (COVID-19), characterized by fever and cough. In diagnostic imaging, the afflicted population showed pathognomonic findings of pneumonia. What started out as an epidemic in China, rapidly spread across geographical locations with a significant daily increase in the number of affected cases. According to the World Health Organization (WHO) reports, the range of worldwide mortality is 3 to 4%. Maternal adaptations and immunological changes predispose pregnant women to a prolonged and severe form of pneumonia, which results in higher rates of maternal, fetal, and neonatal morbidity and mortality. There is limited data about the consequences of COVID-19 in pregnancy, thereby limiting the prevention, counseling, and management of these patients. The objective of this literature review is to explore pregnancy and perinatal outcomes of COVID-19, complications, morbidity, and mortality in this sub-population. We conducted a literature review pertaining to COVID-19 and pregnancy in databases such as: PubMed, Google Scholar, and Science Direct. The studies we chose to focus on were systematic reviews, meta-analysis, case series, and case reports. Twenty four articles were reviewed regarding COVID-19 and pregnancy, complications and their outcomes. Due to immunological changes during pregnancy as evidenced by the flaring of auto-immune diseases; pregnant women may be at an increased risk for infection. Women (19.7%) who had underlying comorbidities such as gestational DM, HTN, hypothyroidism, and autoimmune disease, COPD, or HBV infection were considered high risk. The most common maternal outcomes were premature rupture of membranes (PROM) and pre-eclampsia. Asthma was the most common comorbidity associated with maternal mortality. The most common neonatal complications were fetal distress leading to NICU admissions and preterm birth <37 weeks. The most common laboratory changes were elevated CRP and lymphocytopenia. Most patients underwent C-section due to their underlying comorbidities. Pregnant and lactating women did not shed viral particles through their vaginal mucus and milk, as evidenced by negative nucleic-acid tests of these secretions. Neonatal infections as demonstrated by positive RT-PCR were rare, but direct evidence supporting intrauterine transmission was not confirmed. Direct evidence indicating vertical transmission of COVID-19 is not available, but risk for transmission cannot be ruled out. Pregnant women should be closely monitored due to increased risk of adverse outcomes.

## INTRODUCTION

In December 2019, a novel strain of severe acute respiratory syndrome (SARS-CoV-2), was declared a cause of respiratory illness, called coronavirus 2019 (COVID-19), classically characterized by fever and cough. In diagnostic imaging, the afflicted population exhibited pathognomonic findings of pneumonia. What started out as an epidemic in China, rapidly spread across geographical locations, with a significant daily increase in the number of affected cases. According to the World Health Organization (WHO) reports, the range of worldwide mortality is 3 to 4%.

Pregnancy is a myriad of immunological states. The maternal immune system prepares itself to establish and maintain tolerance to the fetus’s allergenicity, as well as retain the ability to protect the body against microbial invasion. A successful pregnancy relies on finely tuned immune adaptations, both systemically and locally. Maternal immunological states actively adapt and change with the growth and development of a fetus at different gestational stages: pro-inflammatory state (beneficial to the implantation and placentation of the embryo) in the first trimester, anti-inflammatory state (helpful for fetal growth) in the second trimester, and a second proinflammatory state (preparing for the initiation of parturition) in the third trimester [1].

Maternal adaptations and immunological changes during pregnancy tend to predispose pregnant women to a prolonged and severe form of pneumonia, which ultimately results in higher rates of maternal, fetal, and neonatal morbidity and mortality.

Systemic maternal viral infections can also play a factor in affecting the outcome of a pregnancy [2].Previously, studies have shown that SARS infections during pregnancy can lead to high rates of spontaneous abortion, premature birth, and intrauterine growth restriction [3]. However, there has not been evidence of vertical transmission of SARS infections from the mother to the child [3]. Therefore, these pregnancy complications may be caused by the direct effect of the viruses on mothers. Although current evidence is limited, we cannot disregard the potential risks among pregnant women and their fetus. Recent literature indicates that in severe cases, COVID-19 infection is associated with a cytokine-storm, which is characterized by increased plasma concentrations of interleukins 2 (IL-2), IL-7, IL-10, granulocyte-colony stimulating factor, interferon-γ-inducible protein 10, monocyte chemoattractant protein 1, macrophage inflammatory protein 1 alpha, and tumor necrosis factor α (TNF-α) [4], which may be caused by ADE [5]. Since pregnant women in their first and third trimesters are in the pro-inflammatory state, the cytokine-storm induced by SARS-CoV-2 may induce a more severe inflammatory state. Moreover, the occurrence of maternal inflammation as a result of viral infection during pregnancy can affect several aspects of fetal brain development and may lead to a wide range of neuronal dysfunctions and behavioral phenotypes that are unmasked later in postnatal life [6].

The most common symptom at the onset of COVID-19 infection is a fever, which can be associated with an increased risk of attention-deficit/hyperactivity disorder (ADHD) in the offspring during school years.

There is limited data about the consequences of COVID-19 in pregnancy, thereby limiting the prevention, counseling, and management of these patients. The objective of this literature review is to explore pregnancy and perinatal outcomes of COVID-19, complications, morbidity, and mortality in this sub-population.

## METHODS

### Search method and strategy

A literature review was conducted pertaining to COVID-19 and pregnancy in databases such as: PubMed, Google Scholar, and Science Direct from December 2019 through May 31st, 2021. We used specific keywords: “COVID-19,” “Pregnancy,” “SARS-COV-2,” “Pandemic,” “Maternal,” and “Neonates.” The studies we chose to focus on were systematic reviews, meta-analysis, case series, and case reports.

Articles that did not have patient data, and those limited to specific co-morbidities and organ dysfunctions were excluded to avoid selection bias. Therefore, we had twenty four articles for the final review. Selected articles were independently reviewed by two authors. All disagreements were resolved with a discussion between the two authors, or with input from a third independent reviewer and mutually agreed upon by the authors.

Our review included studies from various countries from across the globe. Referencing was done according to guidelines using Endnote. PRISMA guidelines were followed (Fig. 1)

**Fig. 1.**
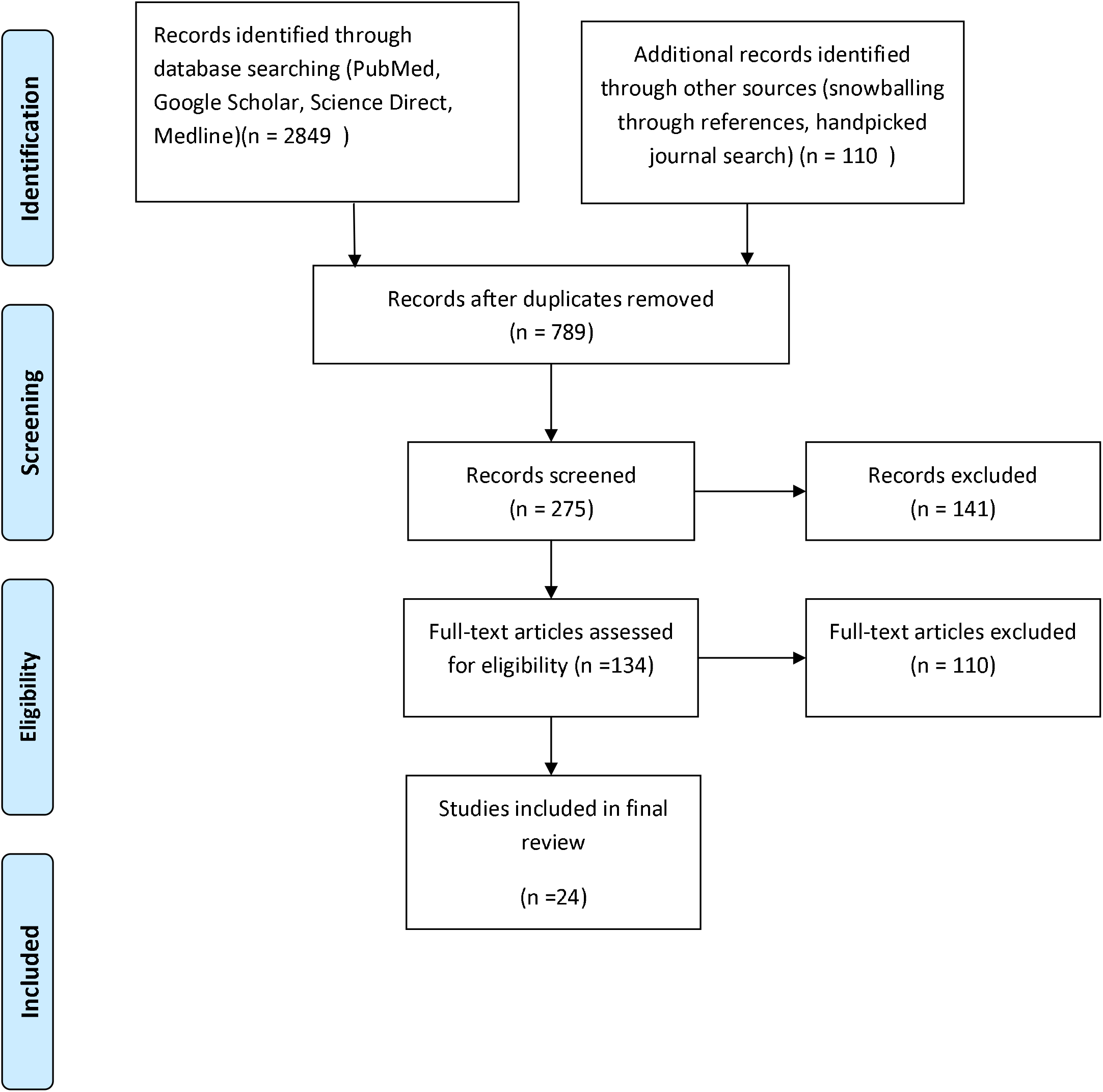
PRISMA diagram for Study design

### Ethical approval and funding

This study did not require ethical approval as data was obtained from already available databases, and patients were not directly involved. No funding was obtained for this review.

## RESULTS

Our results produced a total of 789 studies and after removing duplicates, title, and abstract screening, we chose to focus on 134 studies. A manual search was performed to look for related articles. Twenty four studies were included in the final analysis [Table 1]. Although pregnant women have an immunosuppressed state due to the physiological changes during pregnancy, most patients suffered from mild to moderate COVID-19 pneumonia with no pregnancy loss. Furthermore, this proposes a pattern like the clinical characteristics of COVID-19 pneumonia as compared to that of other adult populations.

**Table 1:**
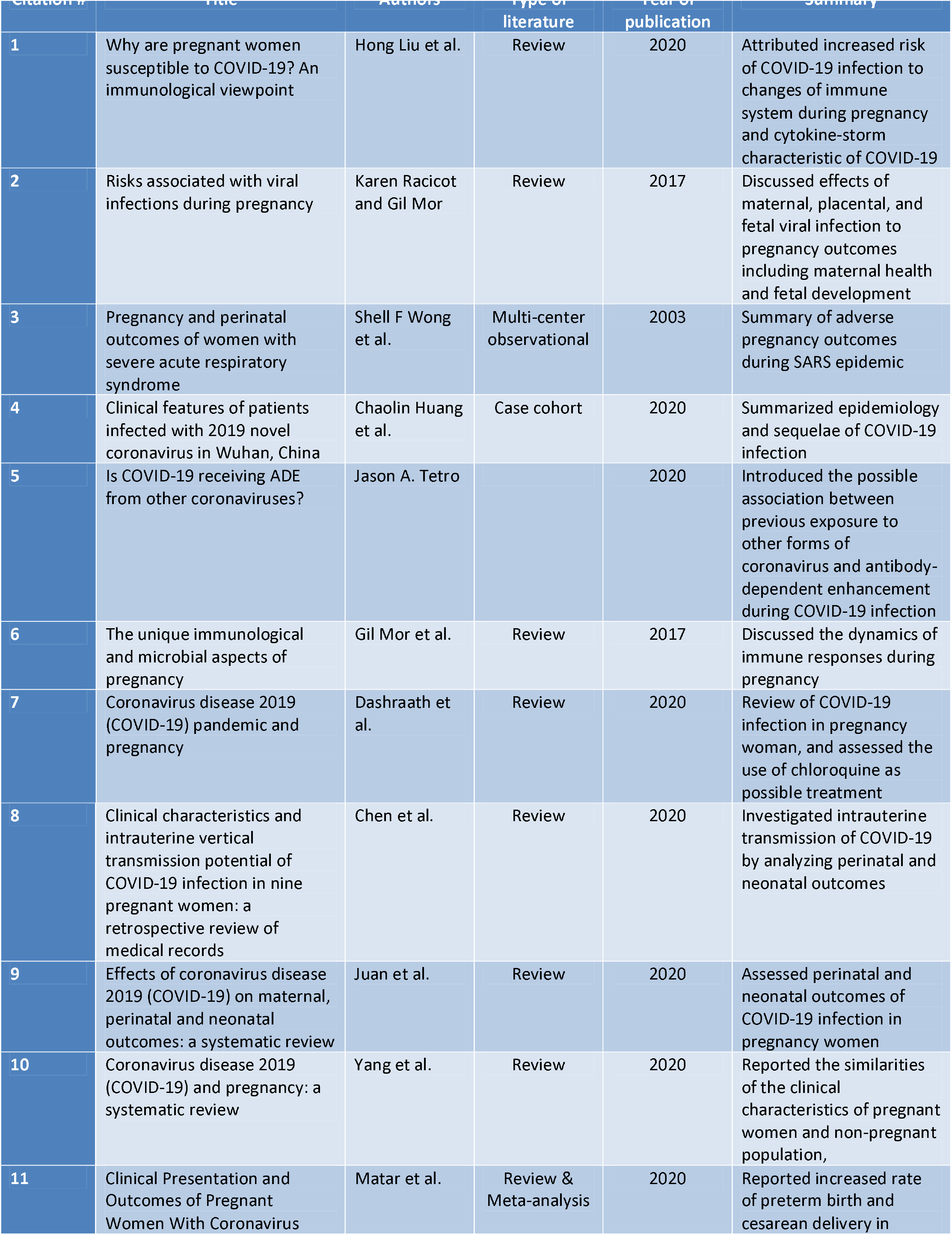
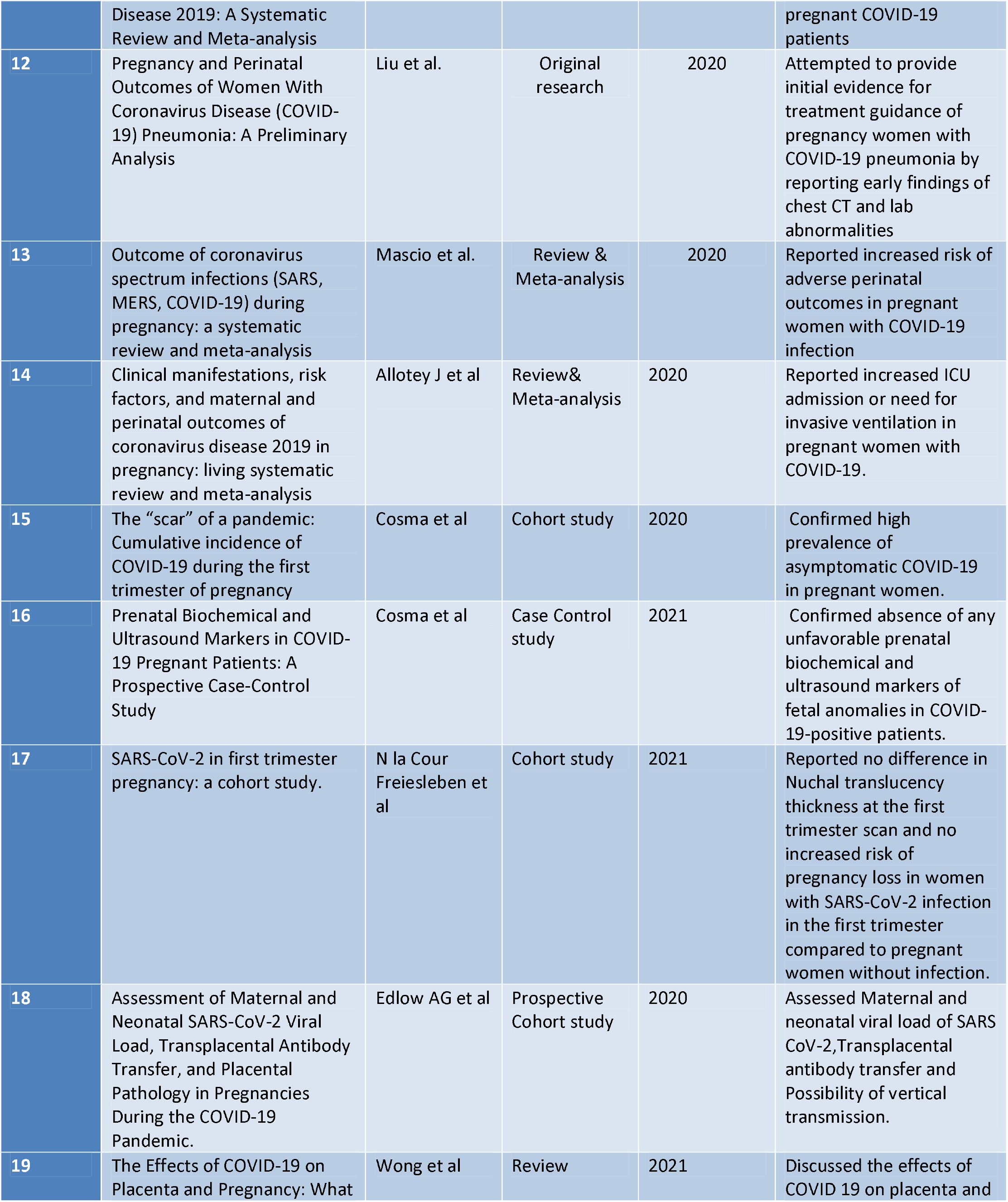
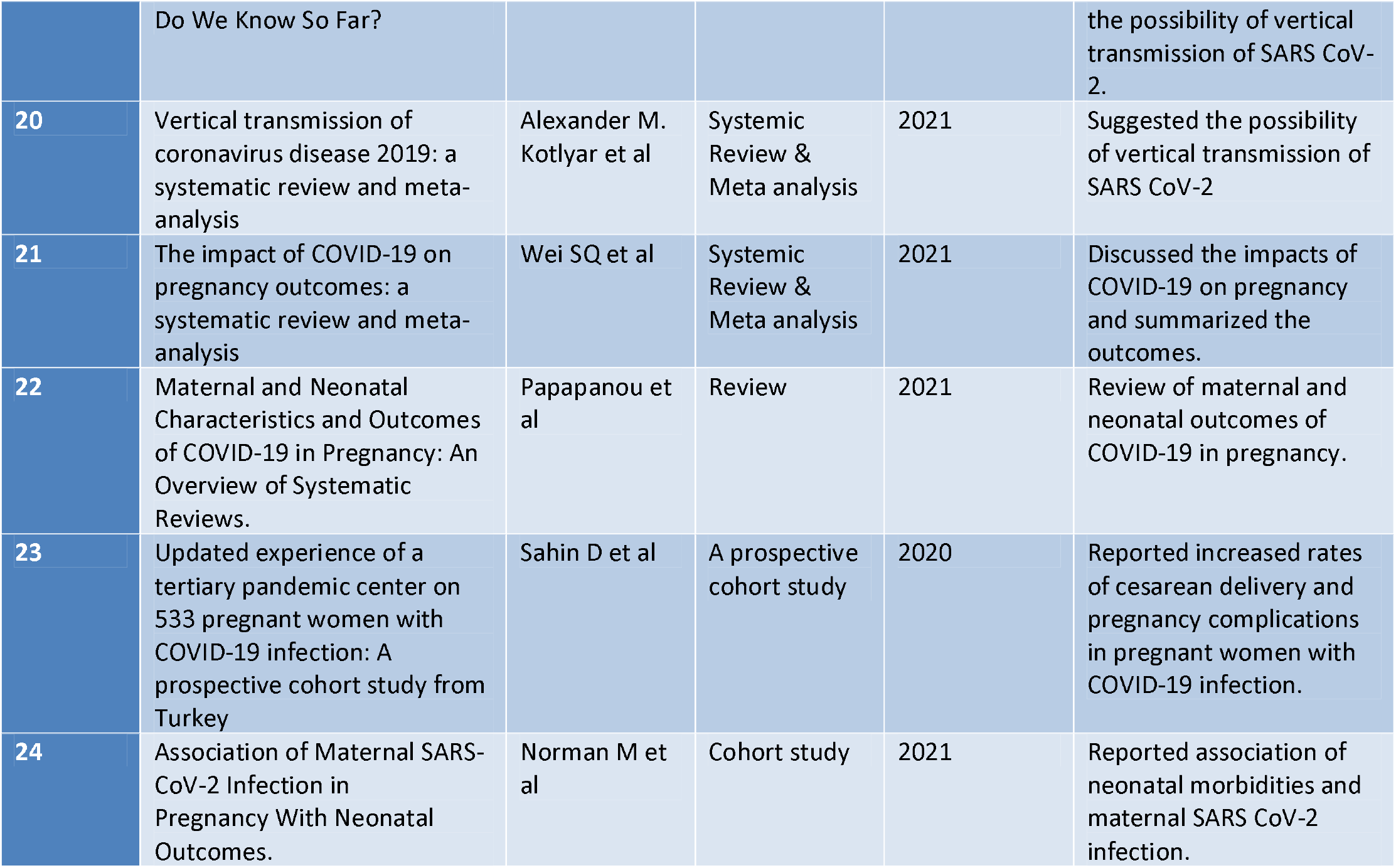
Characteristics of studies

Pregnancy being an immunocompromised state, pregnant women are more at risk of contracting COVID-19. The placenta has been shown to have ACE2 receptors on the cytotrophoblast and syncytiotrophoblast, which is the route through which COVID-19 gains entry. Patients infected with COVID-19 and those who are pregnant are known to have decreased lymphocytes and inhibitory receptors.

Due to immunological changes during pregnancy, as evidenced by autoimmune diseases’ flaring, pregnant women may be at an increased risk for the infection. Most women (19.7%) had underlying comorbidities such as gestational DM, HTN, hypothyroidism, autoimmune disease, COPD, or HBV infection that put them at an increased risk. The most common maternal outcomes were PROM and pre-eclampsia. Asthma was found to be the most common comorbidity associated with maternal mortality. Fetal distress leading to NICU admissions and preterm birth <37 weeks were the neonatal complications. The most common laboratory changes were elevated CRP and lymphocytopenia. The majority of the patients underwent C-section due to their underlying comorbidities. Pregnant and lactating women did not shed viral particles through their vaginal mucus and milk, as evidenced by these secretions’ negative nucleic-acid tests. As demonstrated by positive RT-PCR, neonatal infections were rare, but direct evidence supporting intrauterine transmission was not confirmed.

The findings from this systematic review show that more than 90% of hospitalized pregnant women affected by CoV-SARS infections present radiological signs suggestive for pneumonia, detected either at chest x-ray or computerized tomography (CT) and the most common symptoms are fever, cough, and lymphopenia. Pregnancies affected by CoV-SARS infections have high rates of PTB before 34 and 37 weeks. Preeclampsia and cesarean delivery are also more common than in the general population. The pooled proportion of perinatal mortality is about 10%, while the most common adverse perinatal outcome is fetal distress, with more than half of the newborns admitted in the NICU. Moreover, clinical evidence of vertical transmission was not found in any of the newborns. However, these findings should be interpreted with caution in view of the very small number of included cases and heterogeneity in clinical presentation and perinatal management among the included cases. In a systematic review, women affected by COVID-19 disease had higher rates of preterm birth, and preeclampsia, while the fetus had a 2.4% rate of stillbirth, a 2.4% rate of neonatal death, and higher rate of admission to the NICU. Severe maternal morbidity as a result of COVID-19 and perinatal deaths were reported. Vertical transmission in relation to COVID-19 could not be ruled out.

## DISCUSSION

Pregnancy is an immune compromised state due to several physiologic changes, most commonly cardiorespiratory system changes that place pregnant women even at higher risk of acquiring COVID-19 infection. COVID-19-a newly emerged virus is one of the diseases that can lead to severe consequences; hence a collaborative approach to COVID-19 positive pregnant women is required. It is vital to identify clinical features of SARS-CoV-2 infection in pregnant women as early preventive intervention can lower mortality and morbidity.

The clinical characteristics of pregnant women with COVID-19 resemble those of non-pregnant women. The most common symptoms experienced by these women are fever, cough, dyspnea, fatigue, sore throat, and myalgia [7, 10, 11]. This is supported by a systematic review with 18 studies comprising of 114 pregnant women and determined that (87.5%) of them developed fever, (53.8%) cough followed by (22.5%) having fatigue, (11.3 %) SOB, and (7.5%) sore throat, (16.3%) myalgia {5,6}. 24 studies including 136 pregnant women with confirmed COVID infection were reviewed and found that all of these affected women had similar symptoms as mentioned above [8].However, one of the recent studies showed that pregnant women with covid-19 are less likely to manifest symptoms such as fever, dyspnea, and myalgia, and are more likely to be admitted to the intensive care unit or needing invasive ventilation and extracorporeal membrane oxygenation than non-pregnant women of reproductive age[14]. Increased maternal age, high body mass index, non-white ethnicity, and pre-existing comorbidities were found to be associated with the development of severe disease in pregnancy[14].Moreover, a high prevalence of asymptomatic patients (42.8%) testing positive for SARS Cov 2 further emphasises the need for testing in pregnancy[15].

The most significant blood work abnormalities seen were elevated C-reactive protein in 57 % and lymphocytopenia in 50 % of pregnant women. In addition to this, 81.7 % of all the 136 chest CT scan revealed ground-glass opacity to be the most common atypical imaging findings (9,10,11). Another study done on 15 pregnant women with COVID pneumonia showed Ground-glass opacity with consolidation on Chest CT Scan [10].There are a few studies that highlight the effects of COVID 19 in early pregnancy of which most demonstrated no significant prenatal biochemical or ultrasound markers of fetal anomalies. This was evidenced by a lack of difference in mean nuchal translucency thickness or biochemical markers (pregnancy-associated plasma protein A, alpha-fetoprotein, human chorionic gonadotropin, unconjugated estriol) between cases and controls [16,17].

Although evidence suggesting vertical transmission has not been observed in most reported cases, its possibility cannot be excluded [19]. In a meta-analysis and systemic review including 936 neonates from mothers with coronavirus disease 2019, 27 neonates had a positive result for SARS Cove 2 viral RNA test using nasopharyngeal swab, indicating a pooled proportion of 3.2% (95% confidence interval, 2.2-4.3) for vertical transmission[20].

Low incidence of maternal viremia and non-overlapping placental ACE2 and TMPRSS2 expression were found to be associated with protection against placental infection and vertical transmission in maternal COVID-19.However, reduced trans placental transfer of anti–SARS-CoV-2 antibodies may leave neonates at risk for infection [18].

For maternal related complications, the analysis found that premature rupture of membrane, fetal distress, and stillbirth were the most common complications during pregnancy [12]. In 42 studies involving 548 people who were pregnant, severe COVID-19 infection was found to be strongly associated with preeclampsia, preterm birth, gestational diabetes, and low birth weight when compared with pregnant women having mild COVID 19 infection [21].

When compared with non-pregnant women of reproductive age with covid-19, the odds of admission to the intensive care unit (odds ratio 2.13, 95% confidence interval 1.53 to 2.95; seven studies, 601 108 women) and need for invasive ventilation (2.59, 2.28 to 2.94; six studies, 601 044 women) and ECMO (2.02, 1.22 to 3.34; two studies, 461 936 women) were higher in pregnant and recently pregnant women [14].

Moreover, Pregnancies affected by CoV infections have high rates of PTB before 37 and 34 weeks. Preeclampsia and cesarean delivery are also more common than in the general population. Cesarean delivery was the most common mode of delivery in COVID 19 positive pregnant women [22]. The rate of caesarean delivery was found to be 66.4% in a study involving 533 pregnant women with COVID 19 infection [23].

COVID 19 infection during pregnancy can be associated with adverse neonatal morbidities. This is supported by a prospective cohort study involving 88⍰159 infants from Sweden which showed that some neonatal outcomes (procedures and morbidities) were significantly more common in infants of SARS-CoV-2–positive women (n⍰= ⍰2323) than in infants of comparator women (n⍰= ⍰9275): assisted ventilation at birth (6.4% vs 5.1), intubation at birth (0.6% vs 0.3), admission for neonatal care (11.7% vs 8.4), respiratory distress syndrome (1.2% vs 0.5), use of CPAP (4.9% vs 3.8), mechanical ventilation (1.6% vs 0.5), any respiratory disorder (2.8% vs 2.0%), persistent pulmonary hypertension (0.3% vs 0.1%), antibiotic therapy (2.8% vs 2.0%), and hyperbilirubinemia (3.6% vs 2.5%)[24].

The pooled proportion of perinatal mortality is about 10%, while the most common adverse perinatal outcome is fetal distress, with more than half of the newborns admitted in the NICU. Women affected by COVDID-19 infection had higher rates of preterm birth and preeclampsia, while the babies had a 2.4% rate of stillbirth, 2.4% of neonatal death, and higher NICU admission [13].

## CONCLUSION

COVID-19 being relatively new to the world has made the whole spectrum of population of all ages susceptible due to lack of herd immunity. Pregnancy in general makes women more susceptible to respiratory infections and since COVID predominantly affects lungs, it makes pregnant women a high risk for SARS CoV-2 infection. Due to the pro-inflammatory immune responses in various trimesters of pregnancy by the cytokine storm of COVID-19 infection, pregnant women afflicted with COVID-19 may experience more morbidity and mortality. The maternal infection and inflammatory responses that occur as a response to COVID-19 may affect the fetus even postnatally. With the ever-expanding pandemic, more stringent efforts are required to be made to protect pregnant mothers and growing fetuses. Direct evidence indicating vertical transmission of COVID-19 is not available, but risk for transmission cannot be ruled out. Pregnant women should be closely monitored due to increased risk of adverse outcomes. More studies are required to follow-up the pregnant women having COVID-19 in the first and second trimester to determine pregnancy outcomes and postnatal development of the fetus.

## Data Availability

All authors do not have conflicts of interest and received no funding.

## Notes

### Competing Interest Statement

The authors have declared no competing interest.

### Clinical Trial

N/A

### Funding Statement

All authors did not receive any funding for this paper.

### Author Declarations

There is no direct patient involvement so no requirement of IRB approval.

## REFERENCES

1. Liu, H., Wang, L. L., Zhao, S. J., Kwak-Kim, J et al (2020). Why are pregnant women susceptible to COVID-19? An immunological viewpoint. Journal of reproductive immunology, 139, 103122. https://doi.org/10.1016/j.jri.2020.103122

2. Racicot K., Mor G. Risks associated with viral infections during pregnancy. J. Clin. Invest. 2017;127:1591–1599. doi: 10.1172/JCI87490. [PMC free article] [PubMed] [CrossRef] [Google Scholar]

3. Wong S.F. Pregnancy and perinatal outcomes of women with severe acute respiratory syndrome. Am. J. Obstet. Gynecol. 2004;191:292–297. doi: 10.1016/j.ajog.2003.11.019. [PMC free article][PubMed] [CrossRef] [Google Scholar]

4. Huang C. Clinical features of patients infected with 2019 novel coronavirus in Wuhan, China. Lancet. 2020;395(10223):497–506. doi: 10.1016/S0140-6736(20)30183-5. [PMC free article] [PubMed] [CrossRef] [Google Scholar]

5. Tetro J.A. Is COVID-19 receiving ADE from other coronaviruses? Microbes. Infect. 2020 doi: 10.1016/j.micinf.2020.02.006. Feb 22. pii: S1286-4579(20)30034-4. [PMC free article] [PubMed] [CrossRef] [Google Scholar]

6. Mor G. The unique immunological and microbial aspects of pregnancy. Nat. Rev. Immunol. 2017;17:469–482. doi: 10.1038/nri.2017.64. [PubMed] [CrossRef] [Google Scholar]

7. Dashraath P, Wong JLJ, Lim MXK, Lim LM, Li S, Biswas A et al. Coronavirus disease 2019 (COVID-19) pandemic and pregnancy. Am J Obstet Gynecol. 2020 Jun;222(6):521–531. doi: 10.1016/j.ajog.2020.03.021. Epub 2020 Mar 23. PMID: 32217113; PMCID: PMC7270569.

8. Chen H, Guo J, Wang C, Luo F, Yu X, Zhang W et al. Clinical characteristics and intrauterine vertical transmission potential of COVID-19 infection in nine pregnant women: a retrospective review of medical records. Lancet. 2020 Mar 7;395(10226):809–815. doi: 10.1016/S0140-6736(20)30360-3. Epub 2020 Feb 12. Erratum in: Lancet. 2020 Mar 28;395(10229):1038. Erratum in: Lancet. 2020 Mar 28;395(10229):1038. PMID: 32151335; PMCID: PMC7159281.

9. Juan J, Gil MM, Rong Z, Zhang Y, Yang H, Poon LC. Effect of coronavirus disease 2019 (COVID-19) on maternal, perinatal and neonatal outcome: systematic review. Ultrasound Obstet Gynecol. 2020 Jul;56(1):15–27. doi: 10.1002/uog.22088. PMID: 32430957; PMCID: PMC7276742.

10. Yang Z, Wang M, Zhu Z, Liu Y. Coronavirus disease 2019 (COVID-19) and pregnancy: a systematic review. J Matern Fetal Neonatal Med. 2020 Apr 30:1–4. doi: 10.1080/14767058.2020.1759541. Epub ahead of print. PMID: 32354293.

11. Matar R, Alrahmani L, Monzer N, Debiane LG, Berbari E, Fares J et al. Clinical Presentation and Outcomes of Pregnant Women With Coronavirus Disease 2019: A Systematic Review and Meta-analysis. Clin Infect Dis. 2021 Feb 1;72(3):521–533. doi: 10.1093/cid/ciaa828. PMID: 32575114; PMCID: PMC7337697.

12. Liu D, Li L, Wu X, Zheng D, Wang J, Yang L et al. Pregnancy and Perinatal Outcomes of Women With Coronavirus Disease (COVID-19) Pneumonia: A Preliminary Analysis. AJR Am J Roentgenol. 2020 Jul;215(1):127–132. doi: 10.2214/AJR.20.23072. Epub 2020 Mar 18. Erratum in: AJR Am J Roentgenol. 2020 Jul;215(1):262. PMID: 32186894.

13. Di Mascio D, Khalil A, Saccone G, et al. Outcome of coronavirus spectrum infections (SARS, MERS, COVID-19) during pregnancy: a systematic review and meta-analysis. Am J Obstet Gynecol MFM. 2020;2(2):100107. doi:10.1016/j.ajogmf.2020.100107 https://www.ncbi.nlm.nih.gov/pmc/articles/PMC7104131/ Am J Obstet Gynecol MFM. 2020 Mar 25

14. Allotey J, Stallings E, Bonet M, Yap M, Chatterjee S, Kew T et al. Clinical manifestations, risk factors, and maternal and perinatal outcomes of coronavirus disease 2019 in pregnancy: living systematic review and meta-analysis BMJ 2020; 370: m3320 doi:10.1136/bmj.m3320

15. Cosma, S, Borella, F, Carosso, A, et al. The “scar” of a pandemic: Cumulative incidence of COVID-19 during the first trimester of pregnancy. J Med Virol. 2021; 93:537–540. https://doi.org/10.1002/jmv.26267

16. Cosma, S.; Carosso, A.R.; Borella, F.; Cusato, J.; Bovetti, M. Bevilacqua. et al. Prenatal Biochemical and Ultrasound Markers in COVID-19 Pregnant Patients: A Prospective Case-Control Study. Diagnostics 2021, 11, 398. https://doi.org/10.3390/diagnostics11030398

17. N la Cour Freiesleben, P Egerup, K V R Hviid, E R Severinsen, A M Kolte, D Westergaard, L Fich Olsen et al. SARS-CoV-2 in first trimester pregnancy: a cohort study, Human Reproduction, Volume 36, Issue 1, January 2021, Pages 40–47, https://doi.org/10.1093/humrep/deaa311

18. Edlow AG, Li JZ, Collier AY, et al. Assessment of Maternal and Neonatal SARS-CoV-2 Viral Load, Transplacental Antibody Transfer, and Placental Pathology in Pregnancies During the COVID-19 Pandemic. JAMA Netw Open. 2020;3(12):e2030455. doi:10.1001/jamanetworkopen.2020.30455

19. Wong, Y.P.; Khong, T.Y.; Tan, G.C. The Effects of COVID-19 on Placenta and Pregnancy: What Do We Know So Far? Diagnostics 2021, 11, 94. https://doi.org/10.3390/diagnostics11010094

20. Alexander M. Kotlyar, Olga Grechukhina, Alice Chen, Shota Popkhadze, Alyssa Grimshaw, Oded Tal et al. Vertical transmission of coronavirus disease 2019: a systematic review and meta-analysis, American Journal of Obstetrics and Gynecology,

21. Wei SQ, Bilodeau-Bertrand M, Liu S, Auger N. The impact of COVID-19 on pregnancy outcomes: a systematic review and meta-analysis. CMAJ. 2021 Apr 19;193(16):E540–E548. doi: 10.1503/cmaj.202604. Epub 2021 Mar 19. PMID: 33741725; PMCID: PMC8084555.

22. Papapanou, M.; Papaioannou, M.; Petta, A.; Routsi, E.; Farmaki, M.; Vlahos, N.; Siristatidis, C. Maternal and Neonatal Characteristics and Outcomes of COVID-19 in Pregnancy: An Overview of Systematic Reviews. Int. J. Environ. Res. Public Health 2021, 18, 596. https://doi.org/10.3390/ijerph18020596

23. Sahin D, Tanacan A, Erol SA, Anuk AT, Yetiskin FDY, Keskin HL et al. Updated experience of a tertiary pandemic center on 533 pregnant women with COVID-19 infection: A prospective cohort study from Turkey. Int J Gynaecol Obstet. 2021 Mar;152(3):328–334. doi: 10.1002/ijgo.13460. Epub 2020 Dec 12. PMID: 33131057.

24. Norman M, Navér L, Söderling J, et al. Association of Maternal SARS-CoV-2 Infection in Pregnancy With Neonatal Outcomes. JAMA. Published online April 29, 2021. doi:10.1001/jama.2021.5775

